# LESSONS LEARNED FROM COVID-19 MODELLING EFFORTS FOR POLICY DECISION-MAKING IN LOWER- AND MIDDLE-INCOME COUNTRIES

**DOI:** 10.1101/2024.01.30.24301978

**Authors:** Collins J Owek, Fatuma Guleid, Justinah K Maluni, Joyline Jepkosgei, Vincent Were, So Yoon Sim, Raymond Hutubessy, Brittany L Hagedorn, Jacinta Nzinga, Jacquie N Oliwa

## Abstract

**Introduction:** The COVID-19 pandemic had devastating health and socio-economic effects, partly due to mitigating policy choices. There is little evidence of approaches that guided policy decisions in settings that had limited modelling capacity pre-pandemic. We sought to identify knowledge translation mechanisms, enabling factors, and structures needed to translate modelled evidence to policy decisions effectively.

**Methods:** We utilised convergent mixed methods in a participatory action approach, with quantitative data from a survey and qualitative data from a scoping review, in-depth interviews, and workshop notes. Participants included researchers and policy actors involved in COVID-19 evidence generation and decision-making. They were mostly from lower-and middle-income countries (LMICs) in Africa, Southeast Asia, and Latin America. Quantitative and qualitative data integration occurred during data analysis through triangulation and during reporting in a narrative synthesis.

**Results:** We engaged 147 researchers and 57 policy actors from 28 countries. We found that the strategies required to use modelling evidence effectively include capacity building of modelling expertise and communication, improved data infrastructure, sustained funding, and dedicated knowledge translation platforms. The common knowledge translation mechanisms used during the pandemic included policy briefs, face-to-face debriefings, and dashboards. Some enabling factors for knowledge translation comprised solid relationships and open communication between researchers and policymakers, credibility of researchers, co-production of policy questions, and embedding researchers in policymaking spaces. Barriers included competition among modellers, negative attitude of policymakers towards research, political influences and demand for quick outputs.

**Conclusion:** Our findings led to the co-development of a knowledge translation framework useful in various settings to guide decision-making, especially for public health emergencies. Furthermore, we provide a contextualised understanding of knowledge translation for LMICs during the COVID-19 pandemic. Finally, we share key lessons on how knowledge translation from mathematical modelling complements the broader learning agenda related to pandemic preparedness and long-term investments in evidence-to-policy translation.

**What is already known on this topic:**

- There has been a multitude of modelling frameworks used in diverse ways to advise the various pandemic responses the world over, to an extent not seen before in public health.
- However, it is likely that not all modelling and evidence was adequate, effectively communicated, or used by policymakers.
- This is especially of concern in many LMICs that had strained health systems and resource constraints pre-pandemic.

**What this study adds:**

- The know-do gap is a bottleneck to rapid, effective policy decisions, especially crucial in emergencies.
- As part of pandemic preparedness, it is necessary to have decision support systems in place.
- To ensure this is done well, there is a need to understand how modelling and analytical methods can rapidly be made available and fully integrated into decision-making processes.

**How this study might affect research, practice, or policy:**

- This study contributed to the co-development of a knowledge translation framework that will be useful in building model-to-policy systems that can be adapted for use in various settings.
- We identified mechanisms required to strengthen knowledge translation in LMICs, and this complements the broader learning agenda related to pandemic preparedness and long-term investments in evidence-to-policy translation.

## Introduction

The COVID-19 pandemic is the most defining global health crisis of our times. According to the latest estimates, there have been over 700 million confirmed cases of COVID-19 and just over seven million deaths globally ^1^. Besides directly causing death and disability, the pandemic also disrupted essential health services, putting additional stressors on health systems that were already under strain, especially in lower-and middle-income countries (LMICs) ^2–5^. Policies to curb the spread of COVID-19 negatively affected economic growth and disrupted social services, leading to untold impacts-the pandemic disproportionately affected the most vulnerable ^2^ ^4^ ^6^.

Understanding the magnitude of the effects of the pandemic on health and economic outcomes was essential to developing policies to respond to the crisis. At the pandemic’s beginning, policy decision-makers needed to know the fundamentals of the pathogen and the risk of spread. As it evolved, they needed to understand the incidence, hospitalisation and mortality rates, the effects of various pharmaceutical and non-pharmaceutical interventions and how to allocate resources optimally. As the pandemic subsided, the focus shifted to recovery and long-term impacts ^7^.

Consequently, there was an unprecedented demand for modelling analytics to understand the pandemic and support various mitigation decisions. Compartmental models were commonly used during the pandemic, to monitor individuals as they transition through various infection states (Susceptible-Infectious-Recovered (SIR) and Susceptible-Exposed-Infectious-Recovered (SEIR) models). Agent-based models were also widely used, employing computer simulations to generate a virtual environment where individuals follow defined rules ^8^. However, not all modelling and evidence were likely adequate, effectively communicated, or effectively used by decision-makers ^9–11^.

Despite considerable resources dedicated to research, transferring findings to practice is often a slow, unpredictable process and a bottleneck to rapid, evidence-based policy decisions needed in emergencies ^12^. This may have resulted in missed opportunities, wasted time and effort, and loss of life during the pandemic. It is, therefore, imperative to minimise the knowledge-to-action gap by understanding that knowledge translation processes occur in an environment of diverse evidence sources under uncertainty, with complex social interactions among various stakeholders. Dealing with uncertainties, mainly how to communicate them to decision-makers, is also a significant bottleneck.

Graham’s knowledge-to-action framework (Fig1) has been tested as a model for planning and evaluating knowledge translation strategies ^12^ ^13^. The framework is based on planned action theories. It divides the knowledge-to-action process into two concepts: i) knowledge creation-where the researchers and policy actors generate policy-relevant questions and the modelling approaches to use them) and ii) action-where the modelling evidence is adapted to the local context and used. Guided by Graham’s framework, we set out to identify good practices, enabling factors, and structures needed for the successful creation and use of modelling evidence during the COVID-19 pandemic as a test case for future emergencies.

**Figure 1.**
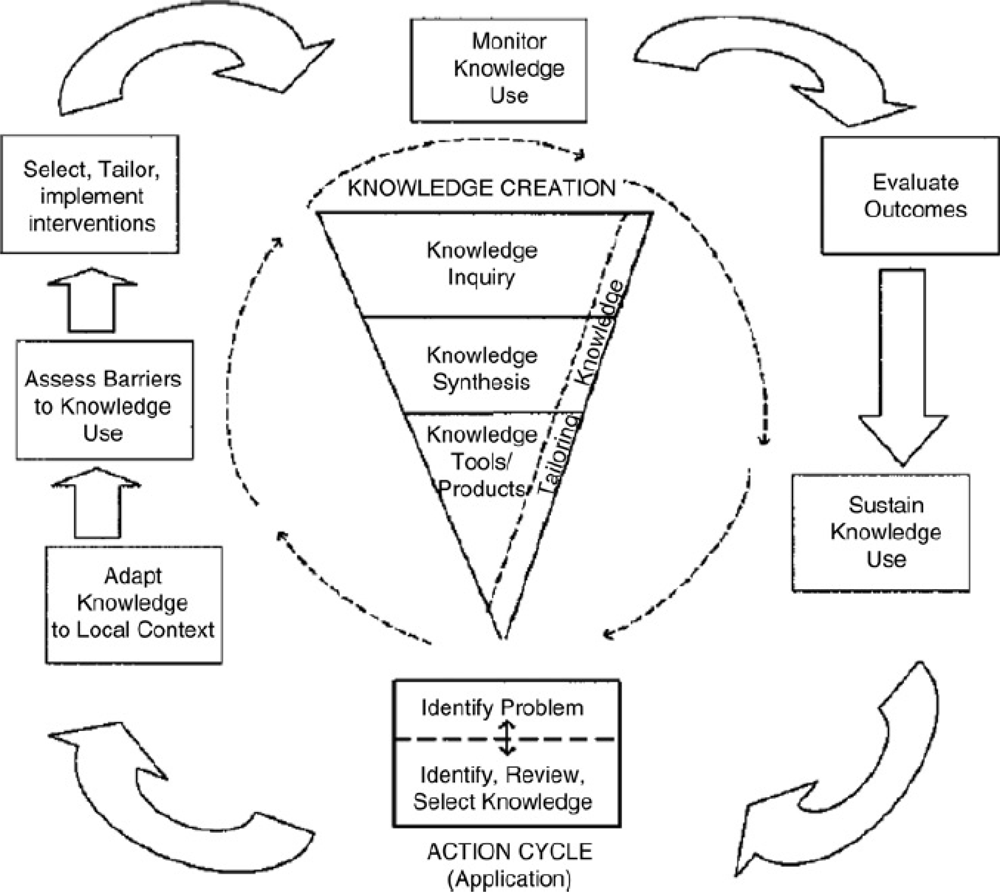
Graham’s Knowledge to Action Framework

We found few studies that explicitly described knowledge translation strategies and how they were used to promote the uptake of modelling evidence for policy decision-making during the pandemic, and most of them were from higher-income settings ^14–16^. LMICs may have had limited modelling and knowledge translation capacity pre-pandemic, and this may have hindered rapid decision-making during the pandemic ^17^. We therefore engaged both researchers and policy actors (primarily drawn from LMICs) to understand how modelling data was used for decision-making during the pandemic, what challenges they faced, and suggestions for improvement. This work resulted in co-creation of a framework to guide evidence-based policy decision-making. It complements the broader learning agenda related to pandemic preparedness and investments in long-term improvement in evidence-to-policy translation.

## Methods

### Study Design

We conducted a mixed methods study in a convergent manner, as described by Creswell and Clark^18^, using participatory action approaches ^19^. Quantitative data were from an online survey, whose insights helped shape qualitative data collection from a scoping review, in-depth interviews, and participant observer notes from learning workshops. Graham’s knowledge-to-action framework helped frame the study objectives, design the initial interview guides, and structure the analysis.

We carried out participatory action research through collective, self-reflective inquiry of local context and social relationships as described by Gilson et al. ^20^. We did this through collaborative and introspective inquiry with participants by looking into their experiences around modelling, knowledge translation and evidence-based policy decision-making during the pandemic. The investigators were thus also participants in the workshops and participated in the co-creation process of the knowledge translation framework. As investigators, we tried to minimise bias by limiting our participation to mainly listening in and only stepping in to facilitate where needed. Our weekly data reflection meetings enabled us to exercise reflexivity to ensure the participants’ perceptions were captured ^21^ ^22^.

### Study Setting and Participants

We purposively selected participants mainly from Africa, Southeast Asia, and Latin America to represent varied levels and capacities of knowledge translation of COVID-19 modelled evidence in lower-resourced settings. Various authors have defined successful knowledge translation as a demonstrable change in knowledge, skill or practice ^23^ ^24^. We determined successful knowledge translation through collective sensemaking with participants on whether and how modelled evidence was considered or used during the public health decision-making process for the pandemic in their respective settings.

We did stakeholder mapping and a scoping review to identify key actors with interest and influence in the COVID-19 modelling knowledge translation space. This helped us generate a database of nearly 200 individuals from 28 countries. Stakeholders included researchers (epidemiologists, infectious disease modellers, economic modellers), with some from the Centres for Epidemiological Modelling and Analysis ^25^ ^26^ and The COVID-19 international Modelling consortium-COMO) ^27^; policy actors/decision makers (government officials, regional and global WHO representatives, task force/technical working group members); the COVID-19 Multi-Model Comparison Collaboration-CMCC ^8^ with members from the World Bank, Health Intervention and Technology Assessment Program-HITAP and WHO Head Quarters; intermediaries (knowledge brokers and boundary organisations); funders; non-governmental organisations; and the public through the media and patient support groups. (S1 Table 1 shows details of the stakeholders, the various countries they were from and the study activities they participated in).

**Table 1.** Sociodemographic characteristics of study participants.

| Characteristic | Survey n (%)<br>N=69 | Interview n (%)<br>N=25 | Workshops n (%)<br>N= 110 |
| --- | --- | --- | --- |
| **Region** |  |  |  |
| Africa | 30 (43%) | 9 (36%) | 60 (55%) |
| Asia | 8 (12%) | 8 (32%) | 16 (15%) |
| Latin America | 10 (15%) | 7 (28%) | 6 (5%) |
| Other | 21 (30%) | 1 (4%) | 28 (25%) |
| **Role played** |  |  |  |
| Researchers/Modellers | 47 (68%) | 17 (68%) | 83 (75%) |
| Policymakers | 22 (32%) | 8 (32%) | 27 (25%) |
| **Years in this role** |  |  |  |
| <5yrs | 20 (29%) | 10 (40%) |  |
| 5-10yrs | 16 (23%) | 12 (48%) |  |
| 11-19yr | 19 (28%) | 2 (8%) |  |
| ≥20yrs | 5 (7%) | 1 (4%) |  |
| Missing | 9 (13%) | 0 |  |
| **Sex** |  |  |  |
| Male | 32 (53%) | 15 (60%) |  |
| Female | 27 (45%) | 10 (40%) |  |
| Other | 1 (2%) | 0 |  |
| **Age group** |  |  |  |
| 25-34yr | 11 (18%) | 3 (12%) |  |
| 35-44yr | 23 (38%) | 6 (24%) |  |
| 45-54yr | 20 (33%) | 14 (56%) |  |
| 55-64yr | 6 (8%) | 2 (8%) |  |
| ≥65yr | 1 (2%) | 0 |  |
| Missing | 7 (10%) | 0 |  |

#### Study Procedures

The study involved a scoping review, an online survey, in-depth interviews and learning workshops as data sources. Fig 2 shows the convergent mixed method approach that we used.

**Figure 2.**
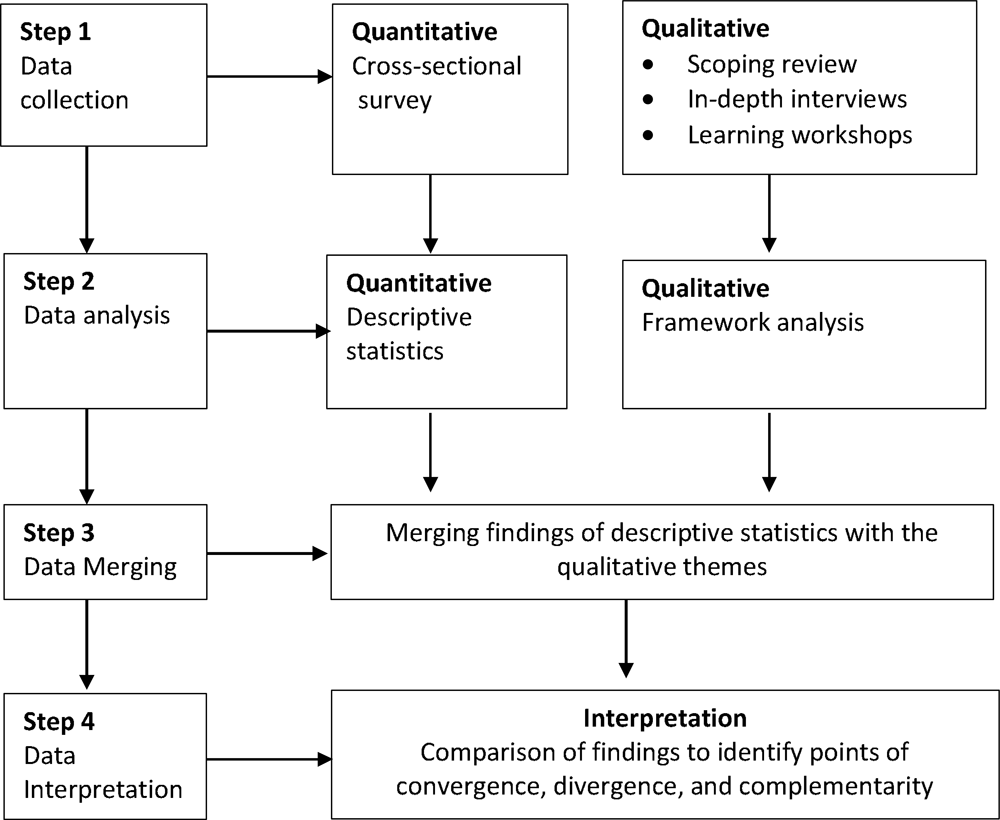
Description of the Mixed Methods Approach

We briefly detail the data collection and analysis processes in the subsequent section.

##### i) Online Survey

We distributed 156 survey questionnaires to potential stakeholders via email in May 2022. We translated and back-translated the surveys in French, Portuguese and Spanish and hosted them on QuestionPro ^28^ (supplementary file S2). We exported data from the survey into RedCAP software and ran consistency checks. We generated descriptive statistics on participant demographics, their knowledge creation and utilisation approaches and the enablers and structures needed for successful knowledge translation.

##### ii) Scoping review

We carried out electronic searches in relevant databases and grey literature in English between March 2020 and April 2022, appraising literature on knowledge translation approaches used to promote the uptake of COVID-19 modelling evidence decision-making.

FG, JM and CO used Rayyan and EndNote to screen titles and abstracts for eligibility. FG and JM did full-text screening and data extraction. Any discrepancies were resolved through discussion with the study team. Supplementary file S3 has details of the search strategy and a summary of the studies.

##### iii) In-depth interviews and learning workshops

We identified key informants focusing on LMIC researchers and policy actors in two stages, initially purposively and then snowballing until the point of saturation. They shared their experiences translating modelled evidence to policy decisions during the pandemic. Graham’s framework initially informed semi-structured interview guides (supplementary file S4) and was later adapted to further explore survey and scoping review findings. We conducted the interviews online due to COVID-19 restrictions between May and December 2022.

Concurrently, we conducted three learning workshops with our stakeholders. The first workshop was to get reflections from participants on their experiences with knowledge translation of modelled COVID-19 data. The second workshop enabled sense-checking and initiated discussions on a knowledge translation guidance framework. The third workshop disseminated further study findings and continued with the co-development of the knowledge translation framework. The investigators were participants in these workshops and took observation notes.

We managed interview transcripts, participant observation notes, and scoping review using NVivo version 12 software. We conducted a thematic framework analysis ^29^ to determine the knowledge creation and utilisation processes used during the pandemic, the participants’ perspectives of what approaches worked well and why, and what could be done differently for future emergencies. CO, JM and JJ read the transcripts and notes and generated initial codes. Discrepancies were discussed and refined during weekly team data reflection meetings. Similar codes were grouped into categories to form a working analytic framework. The process drew on the original research aims guided by Graham’s framework and analytical themes from the recurrence of views or experiences. Next came indexing and charting into a framework matrix. The final stage involved integrating qualitative and quantitative findings from the survey into a narrative synthesis. Anonymised quotes were used for illustrative purposes. Supplementary file S5 has the codebook and S6 snapshot of the framework analysis matrix.

### Ethics approval

This study was approved by The KEMRI (Kenya Medical Research Institute) Scientific and Ethics Review Unit (SERU): Protocol number - KEMRI/SERU/CGMR-C/4425.

## Results

We present the mechanisms used for knowledge translation of modelled evidence for policy decisions during the COVID-19 pandemic in lower-resourced settings, highlighting the enabling factors and infrastructural requirements for successful knowledge translation in future emergencies. We were guided by GRAMMS criteria ^30^ for reporting mixed methods studies, where we illustrated key findings from the quantitative and qualitative arms and how we integrated them.

We engaged 147 researchers and 57 policymakers/advisors from 20 LMICs and eight high-income countries. S1 Tables 1-4 show the details of the countries that the participants were drawn from.

### Participants’ characteristics

Table 1 summarises the sociodemographic characteristics of study participants. The online survey had 69 respondents, 47 (68%) researchers and 22 (32%) policymakers; most (43%) were from Africa. Slightly more than half were male, and nearly 40% were aged 35-44. There were 25 interview respondents, 17 (68%) researchers, and 8 (32%) policymakers, who were distributed across the three main regions (36% from Africa, 32% from Asia, and 28% from Latin America). Most interviewees were male (60%), and more than half were aged 45-54 years. The three learning workshops had 110 participants, 83 (75%) researchers and 27 (25%) policymakers. The workshop attendees were mainly from Africa (55%), Asia (15%) and Latin America (5%).

### Findings from the stakeholder mapping exercise

Figure 3 summarises findings from stakeholder mapping. The stakeholders in category A were high influence but low interest and had to be handled with care/kept informed. They included local policy actors, country office representatives of WHO and local media and patient support groups. Category B was the high-influence and interest group, and we engaged actively with them throughout. They included representatives from members of centres of modelling excellence (CEMA, SACEMA and COMO), the Gates Foundation, regional WHO offices, and CMCC group members (with representation from the World Bank, WHO HQ and HITAP). Category C was the low-influence and low-interest stakeholders that we occasionally consulted mainly to get a developed world perspective, but they were not the focus of our study. This group comprised researchers and partner organisations from high-income settings. Finally, category D had high interest but low influence stakeholders, several of whom needed encouragement to participate. Several stayed engaged throughout. They included researchers, knowledge brokers/intermediaries and task force members from LMICs.

**Figure 3.**
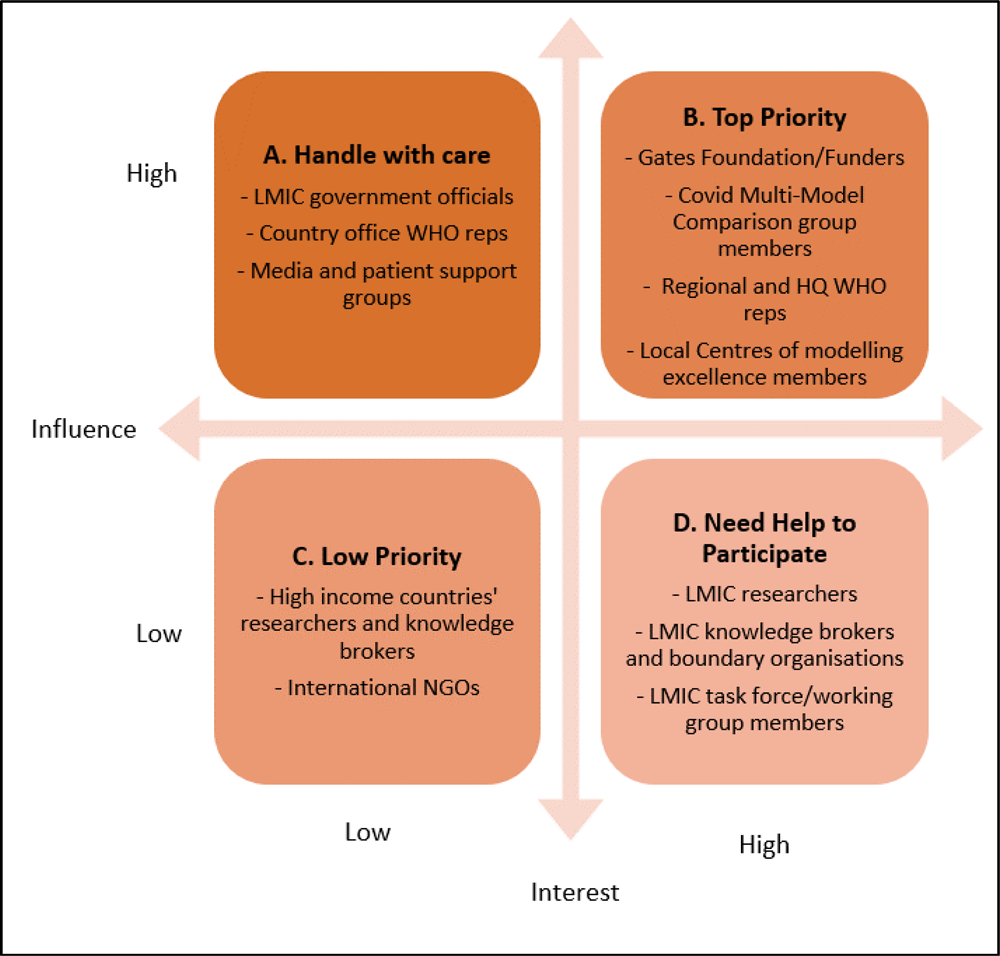
Stakeholder mapping

### Quantitative findings

#### Knowledge creation

We asked survey participants about the knowledge-creation approaches used during the pandemic, see Panel Fig 4 (i). Each proportion represents the number of survey participants from either researchers or policymakers’ groups who responded to each question. Approximately a third of the researchers commented that the policymakers requested modelling data from them, while around a quarter presented their data unsolicited. For the policymakers, almost a quarter reported requesting modelling data, whereas 20% received data unsolicited. Twenty per cent of researchers said they had working relationships with their counterpart policy actors before the pandemic. At the same time, around a quarter reported needing to develop new relationships to respond to the pandemic. Conversely, nearly 30% of policy actors reported having an existing relationship with researchers pre-pandemic, and an equal proportion reported needing to develop new relationships.

**Figure 4.**
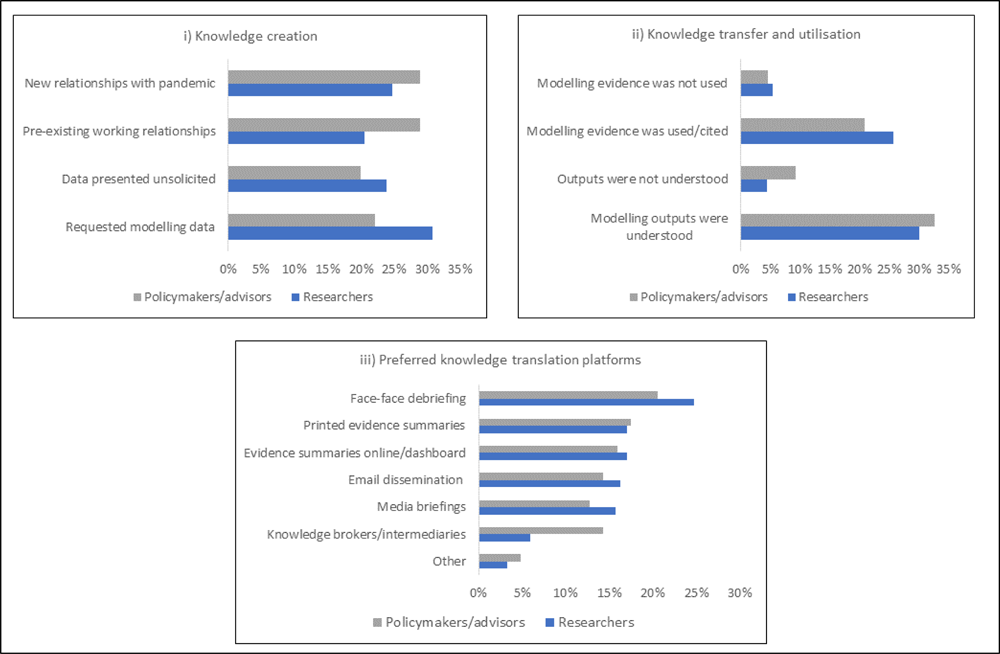
Panel with a summary of responses from survey participants

#### Knowledge transfer/ utilisation

Participants’ responses to the knowledge transfer/utilisation queries are shown in Fig 4 (ii). Around a third of researchers felt that policy actors read and understood their modelling reports. Approximately a quarter responded that the modelling outputs they presented were used in policy decisions, and few perceived that their outputs were not utilised. A third of policymakers reported that they understood the modelled results presented to them, while nearly 10% acknowledged difficulties interpreting the data. A fifth said using model results to guide their decision-making; very few recognised that the data did not influence their policy decisions.

#### Preferred knowledge translation activities

When asked about knowledge translation activities, researchers and policymakers preferred face-to-face debriefing sessions, followed by printed policy briefs of evidence summaries on dashboards (Fig 4 iii). Knowledge brokers/intermediaries were the least preferred platform by both researchers and policymakers.

#### Qualitative findings

The scoping review summarised the knowledge translation strategies used to disseminate modelled evidence during the COVID-19 pandemic. Eight articles were eligible for full-text screening. They were from the WHO Eastern Mediterranean Region ^15^, the ECOWAS region ^16^, New Zealand ^31^, Hungary ^32^, Canada ^33^, Nigeria ^14^, and two had Global representation ^27^ ^34^. All the articles gave descriptive reports on how modelling evidence on COVID-19 was shared with policymakers. Other than information about who was targeted for the knowledge translation strategies, none of the studies provided details about the duration, frequency or timing of events or the personnel and resources required. The specific studies and critical lessons are summarised in supplementary file S3 Table 1.

In the subsequent section, we present emerging themes from the interviews and workshops and triangulate these with findings from the survey and scoping review in a narrative synthesis, highlighting enablers and structures required for effective knowledge translation.

### Enabling Factors for Knowledge Translation

#### Working relationships between modellers and policymakers

Collaborative efforts between researchers (modellers) and policymakers before COVID-19 led to positive working relationships during the pandemic.

> “We’ve spent a lot of time developing relationships with the Ministry of Health and mechanisms for engaging with policymakers…essentially developing prior relationships with policymakers is important because it facilitated knowledge translation during the Covid-19 pandemic.” (Researcher 04_Africa_Interview)

This corroborated survey findings where nearly a third of the researchers and policymakers reported having had pre-existing relationships.

In instances where relationships were poor, there was ineffective knowledge translation where the policy actors were not open to advice. This frequently frustrated the researchers, as was observed in some settings.

> “But the…government didn’t take…any advice from us…The…government ignored completely the advice of…scientists…it was not easy dealing with the policymakers in most regions of X…The government was not very open to the suggestions…” (Researcher_15_Latin America_Interview)

The need for urgent and timely research outputs as the pandemic evolved occasionally led to strained relationships. Running models and packaging outputs required time, yet policymakers expected results quickly, which pressured modellers who worked round the clock to meet the demands.

> “…there’s always some sort of a clash regarding time and expectations between policymakers and researchers. But then how you address those working together over time so that you build those relationships? ” (Researcher 2_Africa_Interview)

In addition, researchers reported the need for some autonomy when building models, even as they acknowledged the need for good relationships and long-term engagement with policymakers before any crisis.

#### Communication

We noted that effective and regular communication between the researchers and the policymakers was fundamental to maintaining relationships. This was substantiated in the scoping review, where open communication was similarly noted as an enabling factor. Good communication ensured that urgent policy questions and research/modelling outputs were promptly exchanged and explained.

> “…it helped a lot to have very regular communication with government… it was really a back and forth, we would come forward with results to present them, and they would ask us for results as well… effective communication with government…was key to… make understanding and the uncertainty a little bit easier…” (Researcher 5_Africa_Interview)

Where there was ineffective communication, researchers sometimes sought alternative channels such as the media, which sometimes forced policy actors to consider advice from researchers due to pressure from the public.

> “…groups were engaging with the media out of frustration, because… they were not able to get through to the policymaking partners…” (Researcher 7_Asia_Interview)

The media, therefore, became an essential channel for information exchange during the pandemic.

> “…we had a lot of work with the press, so we put a lot of effort into giving as many interviews as we can to ask, to reply to the questions that people were having, and also to use the press to spread the recommendations…” (Researcher_15_Latin America_Interview)

#### Trust and Credibility

Continuous engagement and communication created trust between researchers and policymakers, facilitating the uptake of research/modelling outputs if a researcher/institution was perceived to be credible.

> “I think it was also a thing of trust. It was this they knew they could rely on us…because of that relationship, cementing and this constant communication…people know us, and they come to us now…Right now, we get asked for things instead of even one going forward…” (Researcher 5_Africa_Interview)

This was corroborated by workshop participants, one of whom emphasised that building trust takes time.

> “…building trust takes time. It can’t happen overnight during an emergency…” (Researcher_Workshop 1)

Trust and credibility were also key enablers of knowledge translation identified in the scoping review. Likewise, in the survey, most researchers reported that policymakers specifically requested COVID-19 modelling data from them to guide the decision-making, implying trust in their evidence.

#### Co-creation

Where trust and long-standing relationships existed, co-creation of evidence was possible. Co-creation here refers to the engagement of policymakers with researchers in knowledge generation processes, including generation and prioritisation of policy research questions, evidence synthesis, development of models, and interpretation of model outputs. The co-creation process was perceived as a vital enabler in using modelling evidence to inform decisions on COVID-19 response strategies.

> “One of the strategies is coproduction of knowledge…a lot of the evidence generation processes we were… actively involving the policymakers…so that they become part of providing the solution…” Researcher and Policy Advisor_Africa_Interview

We also noted this in the workshops where participants reiterated the importance of co-creation in getting modelled evidence used in policy decision-making.

> “Other organizational-level factors that were important for COVID-19 were organisations that had documented processes for co-creating models or engaging the government from the beginning and model creation. And were much more successful at making sure that their models were…relevant to government priorities and policy needs…” (Researcher, Workshop 1)

Furthermore, research institutions with previous collaborations in the knowledge creation process had established positive working relationships, which made it more likely for their evidence to be considered during the pandemic decision-making.

> “Institution Y has invested in co-creating research with the Ministry of Health, so all the COVID-19 research that we did over the past two years was a collaboration between Institution Y and the Ministry of Health…all our outputs have both Institution Y’s scientists and Ministry of Health employees. and we’ve been invited severally to make presentations to policymakers.” (Researcher 4_Africa_Interview)

In the scoping view, the involvement of policymakers in the generation of evidence encouraged ownership of the process, which enabled them to use the evidence in decision-making. Similarly, in the survey, most policymakers reported working jointly with researchers to develop policy questions for decision-making during the pandemic.

#### Embeddedness

Besides, co-creation, an embedded approach where researchers were situated within policymaking spaces, was an effective way for both researchers and policymakers to be actively involved in generating evidence. *We also identified some policymakers attached to research organisations to participate in knowledge generation.* The participation of policymakers allowed them to have a better understanding of knowledge generation processes. Thus, they were better positioned to interpret and potentially use model outputs for decision-making.

> “…an embedded approach … we adopted a way of working with policymakers…we do that research within policymaking spaces and with active involvement and participation of policymakers…that has evolved over time and because of that it has really helped to facilitate the actual application of the research that we do.” (Researcher 2_Africa_Interview)

Sometimes, researchers were invited to present their findings during live policy discussions. This was perceived to encourage the uptake of recommendations and their implementation.

> “….so that allows the scientists to come and attend those cabinet sessions if at all there is an issue on the agenda…is one of the greatest ways to have this evidence to the policymakers…” (Policymaker 3_Africa_Interview)

### Structures needed for successful knowledge translation

#### Capacity building

We identified capacity building for local modelling expertise as critical for successful knowledge translation. Researchers and policymakers both underscored the importance of having several modellers who could generate context-relevant models for policy decision-making.

“We have shortage of modelling expertise…policymakers wanted to have geographic specific interventions for Covid-19 response…we as a country, kept waiting for [researchers]to be able to guide but also inform us of which model and what is likely to work out where.” (Policymaker 03, Africa_Interview)

Workshop participants brought up that having regional centres of excellence responsible for training modellers, fundraising, building collaborations, and forming stable links with policymakers for co-creation purposes would be a good aspirational goal.

Beyond technical modelling capacity strengthening, the participants identified a need to train researchers and policymakers in science communication, specifically in disseminating scientific outputs to a lay audience. Several researchers found themselves in the deep end during the pandemic, having to communicate their findings with no prior training.

> “…it was hard, but we really didn’t have any previous training on how to do that…we had this big group, and we saw which were the good speakers, the people that had more clarity to spread the ideas, and then we just pushed them to do the interviews… ”(Researcher 15_Latin America_Interview)

Workshop participants also emphasised that policymakers needed training to empower them to interpret and use scientific evidence for decision-making, including understanding the uncertainties in modelling and which questions can or cannot be answered by modelling data.

#### Data infrastructure

In addition to capacity building, the participants reported the need for local data systems and policies that made high-quality data available and enabled data sharing. Such infrastructure would allow modellers to develop timely models based on local data and with the capacity to inform local decision-making.

> “…important factors included the availability of high-quality local data and information systems that modellers could quickly pull up and use to develop models. It was helpful when these data systems were transparent and… formatted in a way that made them accessible and usable for modellers.” (Researcher, Workshop 2)

The need for collaboration between academic institutions and the government was underscored to attain better data infrastructure, more so in low-resource settings.

> “…better data systems…improved surveillance, more open data, accessible data and then the other thing is more opportunities for collaboration between academia and government because you have to build trust.” (Researcher 14_Latin America_Interview)

#### Dedicated funding streams

Resources are needed to build capacity for technical modelling, knowledge translation expertise, and data infrastructure. Respondents mentioned political buy-in as critical for governments to invest domestic public resources in modelling and for policymakers to use model outputs for decision-making. Therefore, researchers must build alliances within various levels of government that may lead to dedicated resource allocation for modelling.

> “… getting that political buy-in and the government to invest dedicated resources for modelling to respond to pandemics. I think it is a big challenge in low-and middle-income countries. If you look at countries in the West, most countries have a dedicated resource, a dedicated unit, or a dedicated university who have been assigned the responsibility and also the dedicated resources for those groups” (Researcher 11_Southeast Asia_Interview)

### Framework to guide the use of modelling data to guide policy decision-making

We presented the empirical evidence from our study during the learning workshops. We asked participants to deliberate on how best to support using models for policy decision-making in preparation for future pandemics, especially in countries with limited capacity. We co-developed a framework with an implementation matrix (fig 5) and a simplified road map for policy consumption, whose details are described further in this policy brief ^35^..and publication.

**Figure 5.**
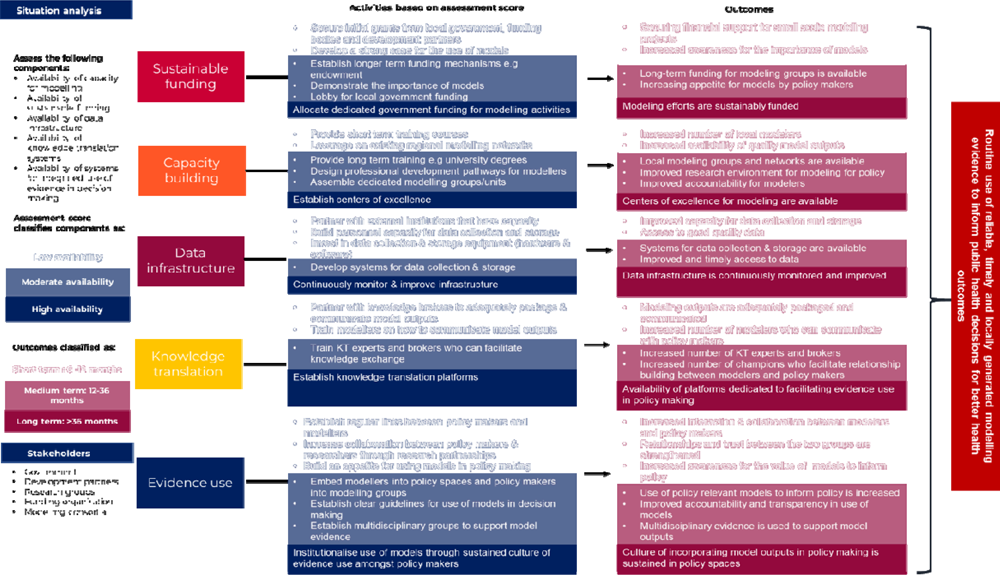
Framework to guide the use of models for Policy decision-making

In summary, the overall goal of the framework is to ensure the routine use of reliable, timely and locally generated modelling evidence to inform public health decisions for better population health outcomes. The final five key components include i) Sustainable funding to support modelling for policy; ii) capacity building leading to centres of excellence for modelling; iii) sustainable systems and structures for the generation and use of model outputs; iv) dedicated platforms for knowledge translation; and v) policymakers engaged throughout the knowledge generation process and thus committed to a culture of using evidence to guide decision making.

## Discussion

Our study led us to understand better the preferred mechanisms used for knowledge translation of modelled data during the COVID-19 pandemic, enabling factors and infrastructural requirements in preparation for future global emergencies, especially in lower and middle-income country settings.

The commonly used knowledge translation activities were face-to-face debriefs, policy briefs, and evidence summaries on dashboards. Relatedly, in studies from humanitarian emergencies, policymakers preferred research findings presented in short messages with key actions highlighted in bullet points or infographics on websites they could easily access and use ^36^. A study in the Mediterranean region likewise described the benefits of sharing research findings via non-technical audio-visual presentations and policy briefs ^15^. Surprisingly, we found that using knowledge brokers/intermediaries was the least popular knowledge translation mechanism; policymakers in LMICs preferred direct engagement with researchers. This is contrary to what was observed from studies from higher-income settings where knowledge brokers promoted a culture of using evidence for decision-making and thought to be an optimal knowledge translation and exchange strategy ^37^ ^38^.

As we explored enablers for effective knowledge translation of modelled evidence during the pandemic, pre-existing relationships between the researchers and policymakers, coupled with open communication, trust, and credibility, were vital to co-creating policy-relevant knowledge products. In examples from Israel, Switzerland, Germany, Canada and South Korea, countries where the government collaborated well with scientists seemed to have better patient outcomes during the pandemic, compared to Italy, Spain, Brazil and the United States, where the government and scientists had more strained relationships with devastating consequences ^32^ ^39–42^. An example from LMICs, Nigeria, had a co-production approach through a presidential taskforce of decision-makers and multi-disciplinary academics. This enabled swift production and effective utilisation of scientific data in response to the COVID-19 pandemic ^14^. Kenya and South Africa similarly had participatory approaches of co-production and embeddedness, where researchers worked alongside policymakers to support collaborative research and learning processes pre-pandemic, which proved helpful in the pandemic responses ^20^.

Barriers to effective knowledge translation included competition among modellers, negative attitude of policymakers towards research, political influences and demand for timeliness of research outputs for policy decisions. Policymakers sometimes cited difficulties in determining the quality of evidence, which has been reported in other studies ^43^. Some said that close integration caused a loss of autonomy and the impact of power dynamics on model quality ^14^. Findings from a review cited lack of time, limited access to research evidence, limited capacity to appraise and translate research evidence and resistance to change as some of the barriers to evidence-informed decision-making ^37^.

Finally, we used participatory approaches to identify and prioritise structures required to navigate the barriers and support the effective translation of modelled evidence to policy, especially in lower-resourced settings. We packaged these in a framework and roadmap to guide policymakers that are described in detail elsewhere ^35^. A need for capacity building particularly stood out. Lower-resourced countries relied on collaborative efforts to cope during the pandemic. This was seen in action through efforts of groups like the COVID-19 Modelling Consortium (COMO)^27^ and the emergence of centres of excellence like SACEMA in South Africa ^26^ and The Centre for Epidemiological Modelling and Analysis (CEMA) in Kenya ^25^. The pandemic also saw work from the COVID-19 Multi-Model Comparison Group (CMMC) that provided guidance to ensure models were relevant, robust and useful for policy decision-making ^34^ ^44^.

### Strengths and Limitations

We used mixed methods, including an online survey, a scoping review, interviews and learning workshops that gave us rich insights into knowledge translation mechanisms, what worked well, where and why, and how best to improve. Furthermore, the study findings contributed to the co-development of a knowledge translation roadmap and framework with an implementation matrix.

The fundamental limitation is that the study happened during the pandemic, which limited interviews to online platforms. We strengthened the quality of our outputs by using mixed methods that enabled us to triangulate our findings, and when restrictions eased, we had an in-person learning workshop.

## Conclusion

The findings from this study led to the co-development of a knowledge translation framework that will be useful in integrating model-policy translation dynamics. The framework can be adapted in various settings to guide decision-making in preparation for and response to public health emergencies. Furthermore, we provide a contextualised understanding of knowledge translation for lower and middle-income countries during the COVID-19 pandemic. Face-to-face debriefings were the most preferred knowledge translation interventions. The critical enabling factor was pre-existing relationships between researchers and decision-makers. In addition, co-creation and embeddedness contributed to successful knowledge translation. Challenges identified included competition among modellers, the negative attitude of some policymakers towards science and political influence. Finally, we provide vital lessons on how knowledge translation from mathematical modelling complements the broader learning agenda related to pandemic preparedness and long-term investments in evidence-to-policy translation.

## Key for supplementary files

S1 Study Participants

S2 Online survey tool

S3 Scoping review

S4 Interview guides

S5 Coding Framework

S6 Thematic Analytic Framework Matrix charting

## Data Availability

The survey responses and qualitative interview transcripts are housed on secure servers at the KEMRI-Wellcome Trust Research Programme. Access applications can be made through the Data Governance Committee, with details available at www.kemri-wellcome.org or email to.

## Acknowledgements

We would like to thank the study participants and all the stakeholders for generously sharing their time and information with this project. We also acknowledge the tireless efforts of our administrative team led by Metrine Saisi and Elizabeth Isinde, that helped coordinate the smooth running of our workshops with participants coming in from all over the world.

The contents of this article are solely the responsibility of the authors and do not represent the official views of WHO or the Gates Foundation.

## Authorship Contributions

JNO and BLH conceptualised the idea of the study. CJO, JM and JJ were responsible for primary data collection. CJO, JM, JJ, FG and JNO met weekly for data reflection and analysis. JN, VW, SSY, and RH joined the rest of the study team monthly to brainstorm on the themes from the data to refine the framework, working on feedback from the other contributors. CJO and JNO wrote the early drafts of the manuscript and refined it with feedback and input from all the authors. All authors approved the final version of the manuscript.

## Funding

Funds from the Bill and Melinda Gates Foundation grant (INV-034291) awarded to Dr Jacquie N Oliwa supported this work. The views in this article are those of the authors and do not represent the official views of the Gates Foundation or WHO. The funders had no direct role in study design, data collection and analysis, the decision to publish or the preparation of the manuscript.

## Competing interests

The authors completed the ICMJE Disclosure of Interest Form (available upon request from the corresponding author) and some have declared technical consultancies and travel support outside the submitted work. No commercial companies were involved.

## Patient and public involvement

Patients and the public were not involved in this research’s design, conduct, reporting, or dissemination plans.

## Patient consent for publication

Not required.

## Supplemental material

### S1 Study Participants

**Table 1.** Countries that participated in the study.

| Country | Region | Country<br>Classification<br>(World<br>Bank) | Survey | Interviews | Learning<br>Workshops |
| --- | --- | --- | --- | --- | --- |
| 1. Kenya | Africa | LMIC | X | X | X |
| 2. South Africa |  | UMIC | X | X | X |
| 3. Ghana |  | LMIC | X |  |  |
| 4. Congo |  | LMIC | X |  |  |
| 5. Nigeria |  | LMIC | X |  | X |
| 6. Rwanda |  | LIC | X |  | X |
| 7. Uganda |  | LIC | X | X | X |
| 8. Tunisia |  | LMIC |  | X | X |
| 9. Thailand | Asia | UMIC | X | X | X |
| 10. India |  | LMIC | X |  |  |
| 11. Malaysia |  | UMIC | X | X | X |
| 12. Singapore |  | HIC | X | X | X |
| 13. Kyrgyzstan |  | LMIC | X | X |  |
| 14. Argentina | Latin America | UMIC | X | X |  |
| 15. Brazil |  | UMIC | X | X |  |
| 16. Panama |  | HIC | X |  |  |
| 17. Colombia |  | UMIC | X |  |  |
| 18. Ecuador |  | UMIC | X |  |  |
| 19. Chile |  | HIC | X | X |  |
| 20. Paraguay |  | UMIC | X |  |  |
| 21. UK | Other regions | HIC | X |  | X |
| 22. USA |  | HIC | X |  | X |
| 23. Switzerland |  | HIC | X |  | X |
| 24. Australia |  | HIC | X |  | X |
| 25. France |  | HIC |  |  | X |
| 26. Netherlands |  | HIC |  |  | X |
| 27. New Zealand |  | HIC |  |  | X |
| 28. Canada |  | HIC |  |  | X |

**Table 2.** Survey Respondents.

| Region | Country | Researchers | Policymakers | Total |
| --- | --- | --- | --- | --- |
| **Africa** | Kenya | 11 | 8 | **19** |
|  | South Africa | 2 | 2 | **4** |
|  | Multiple Countries | 1 | 1 | **2** |
|  | Ghana | 0 | 1 | **1** |
|  | Congo | 0 | 1 | **1** |
|  | Nigeria | 1 | 0 | **1** |
|  | Rwanda | 1 | 0 | **1** |
|  | Uganda | 1 | 0 | **1** |
| **Southeast Asia** | Thailand | 1 | 2 | 3 |
|  | India | 0 | 1 | 1 |
|  | Malaysia | 1 | 0 | 1 |
|  | Singapore | 3 | 0 | 3 |
| **Latin America** | Argentina | 2 | 0 | 2 |
|  | Brazil | 1 | 0 | 1 |
|  | Panama | 1 | 0 | 1 |
|  | Colombia | 2 | 1 | 3 |
|  | Ecuador | 1 | 0 | 1 |
|  | Chile | 1 | 0 | 1 |
|  | Paraguay | 1 | 0 | 1 |
| **Others** | UK | 10 | 1 | 11 |
|  | Multiple | 2 | 0 | 2 |
|  | USA | 2 | 2 | 4 |
|  | Switzerland | 0 | 2 | 2 |
|  | Kyrgyzstan | 1 | 0 | 1 |
|  | Australia | 1 | 0 | 1 |
| **TOTAL** |  | **47** | **22** | **69** |

**Table 3.** Interviews Respondents.

| **Region** | **Country** | **Researchers** | **Policymakers** | **Total** |
| --- | --- | --- | --- | --- |
| **Africa** | Kenya | 4 | 1 | 5 |
|  | South Africa | 1 | 0 | 1 |
|  | Multiple Countries | 0 | 1 | 1 |
|  | Uganda | 0 | 1 | 1 |
|  | Tunisia | 1 | 0 | 1 |
| **Southeast Asia** | Thailand | 4 | 1 | 4 |
|  | Malaysia | 2 | 0 | 2 |
|  | Singapore | 1 | 0 | 1 |
| Latin America | Argentina | 0 | 1 | 1 |
|  | Brazil | 2 | 3 | 5 |
|  | Chile | 1 | 0 | 1 |
| Others | Kyrgyzstan | 1 | 0 | 1 |
| **TOTAL** |  | **17** | **8** | **25** |

**Table 4.**
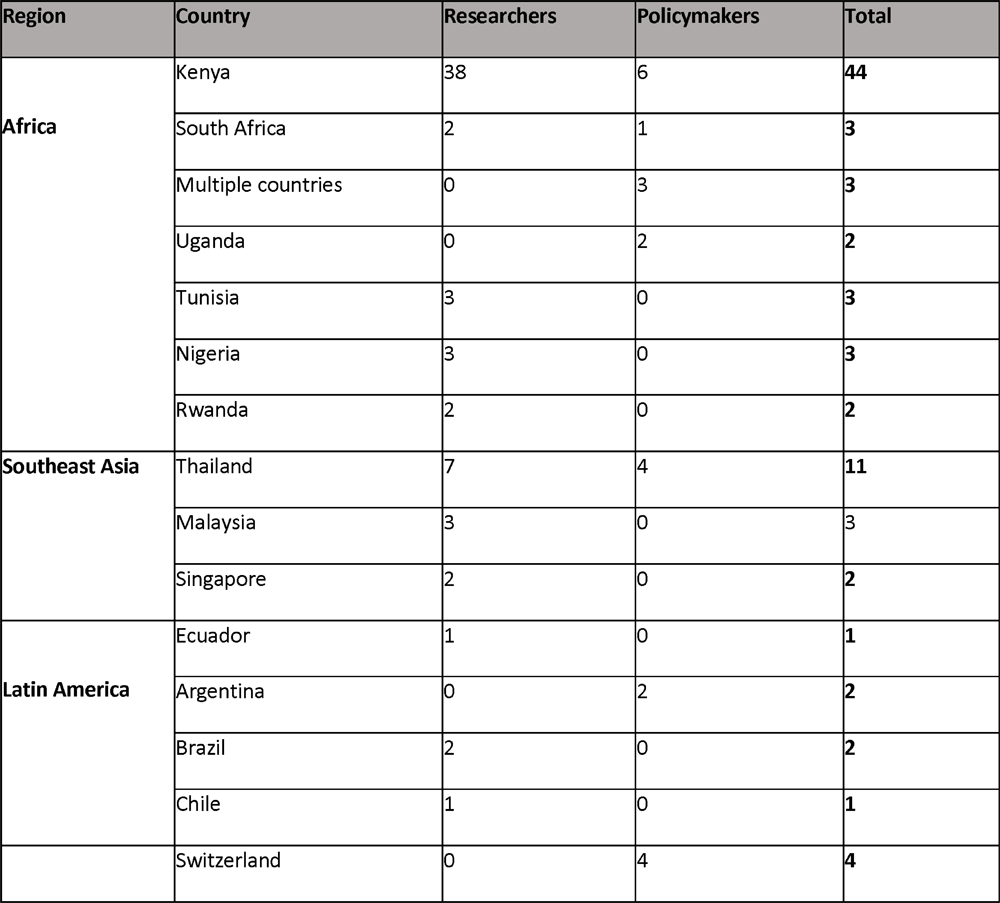
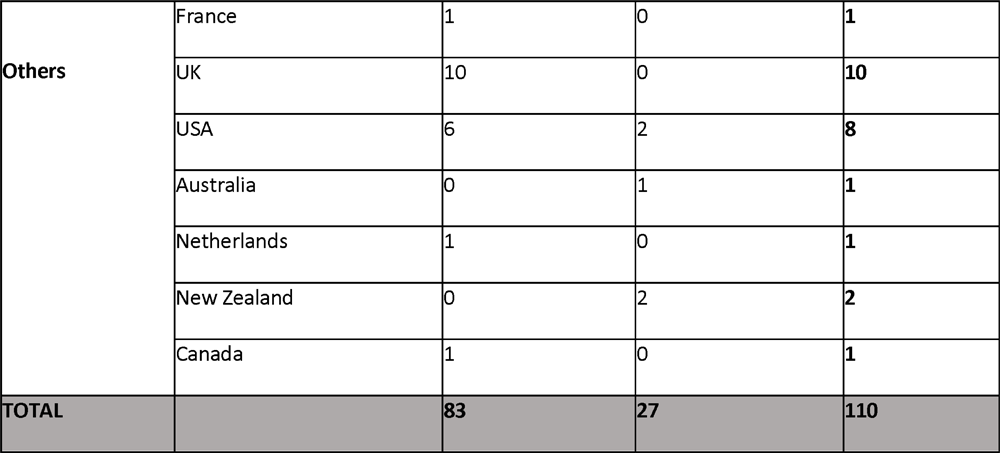
Learning workshop participants.

### S2 Online Survey tool (English)

This section is to collect general information about you and the professional role (s) that you play. 1. Indicate your gender

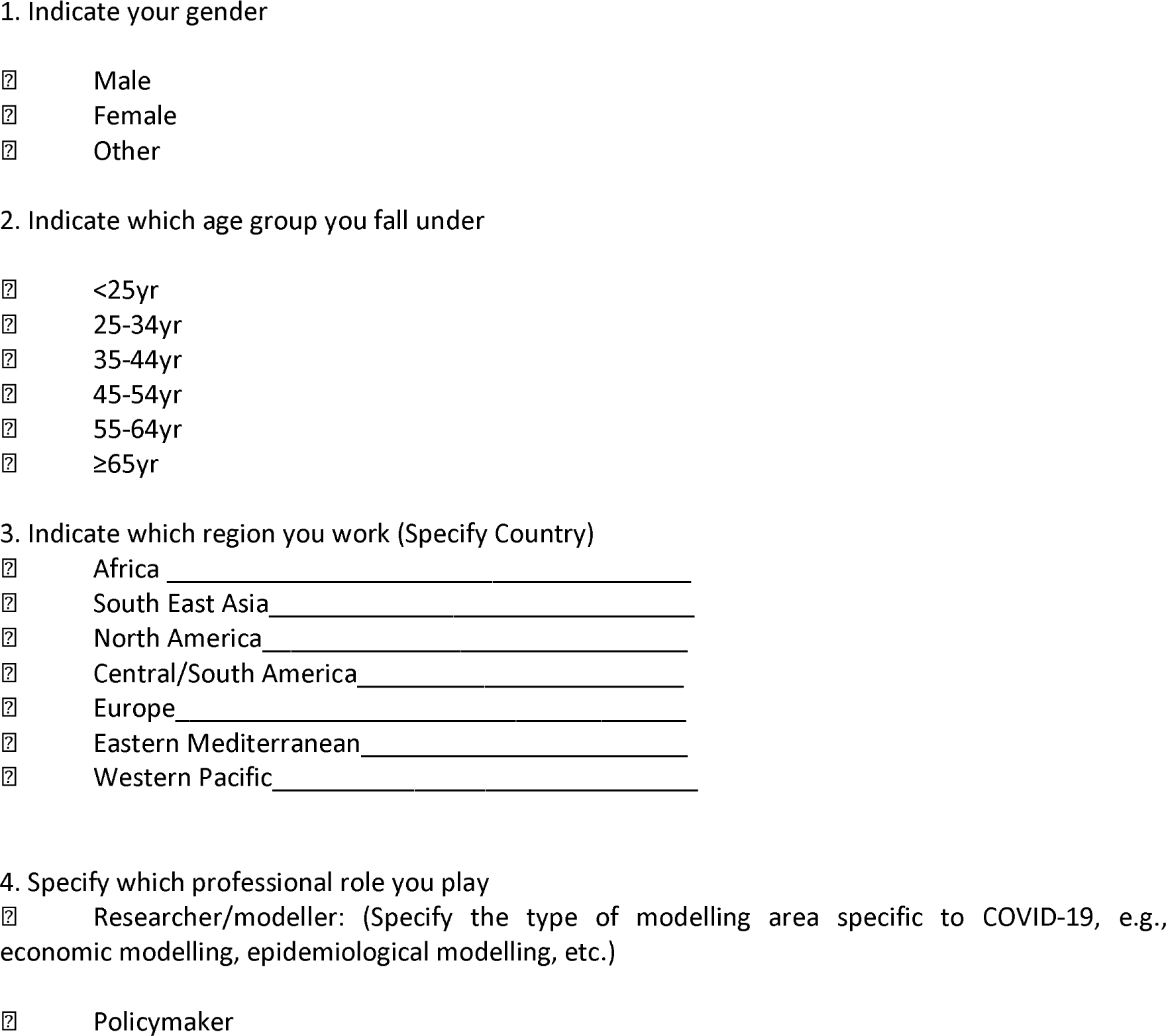
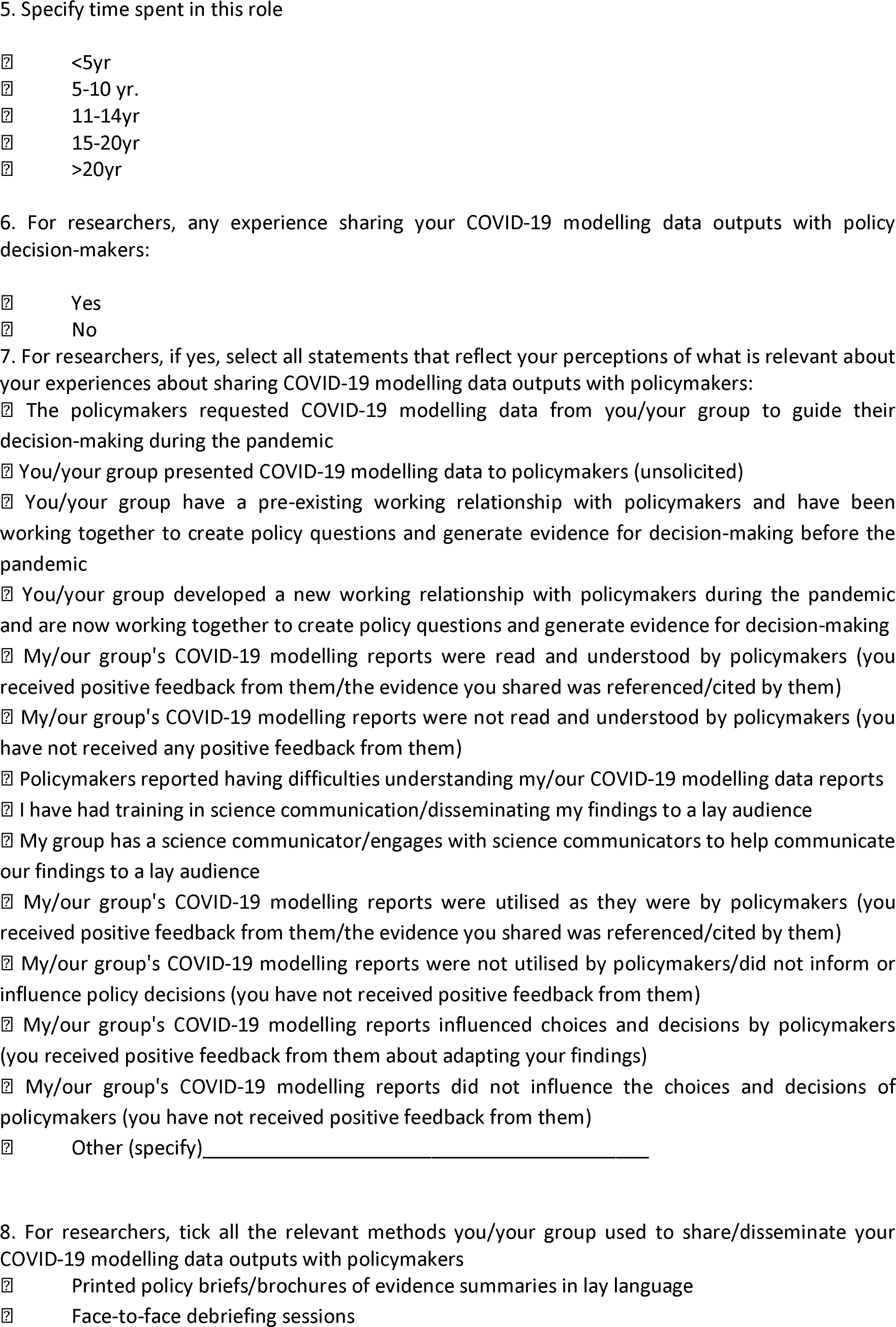
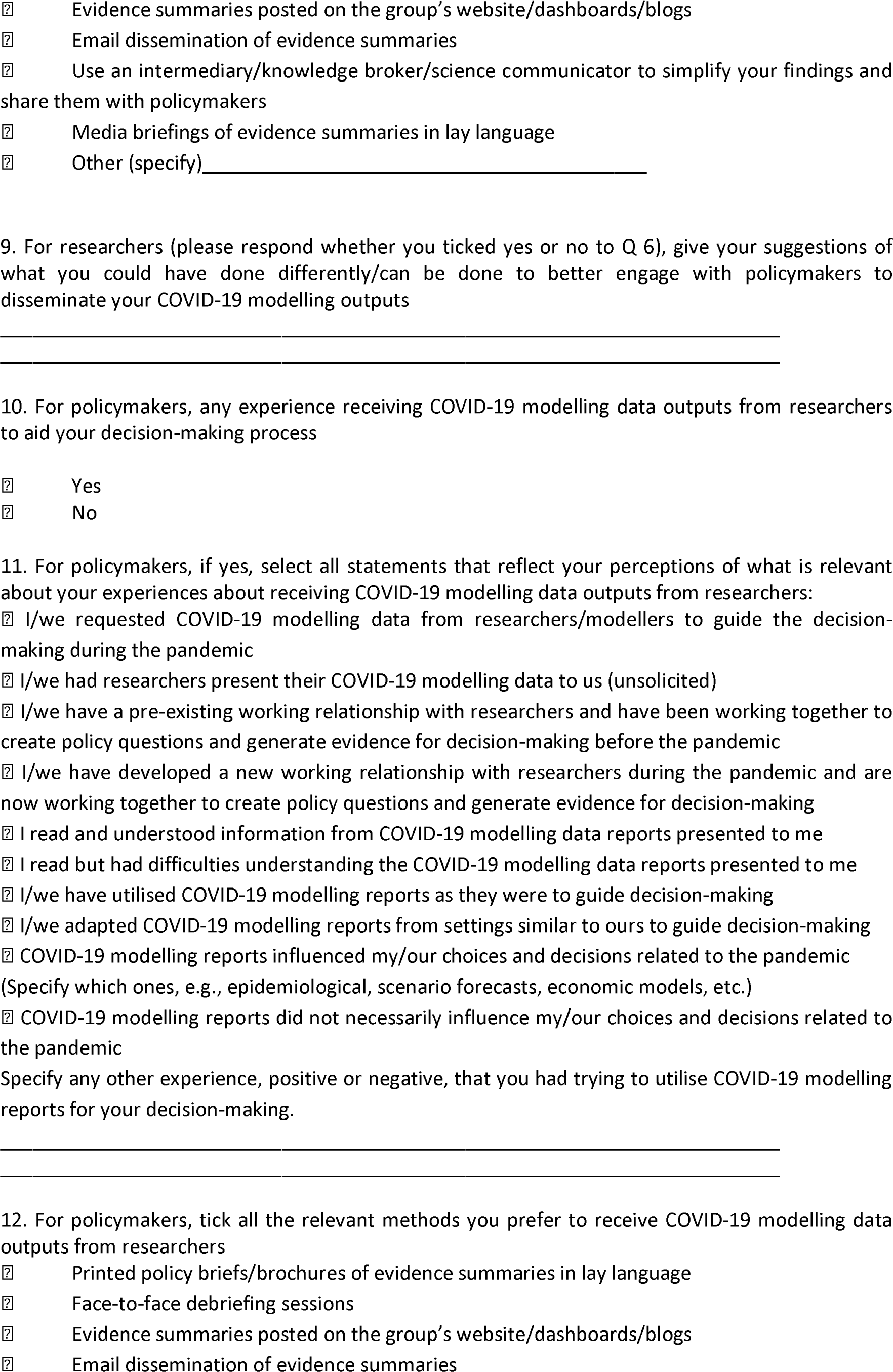
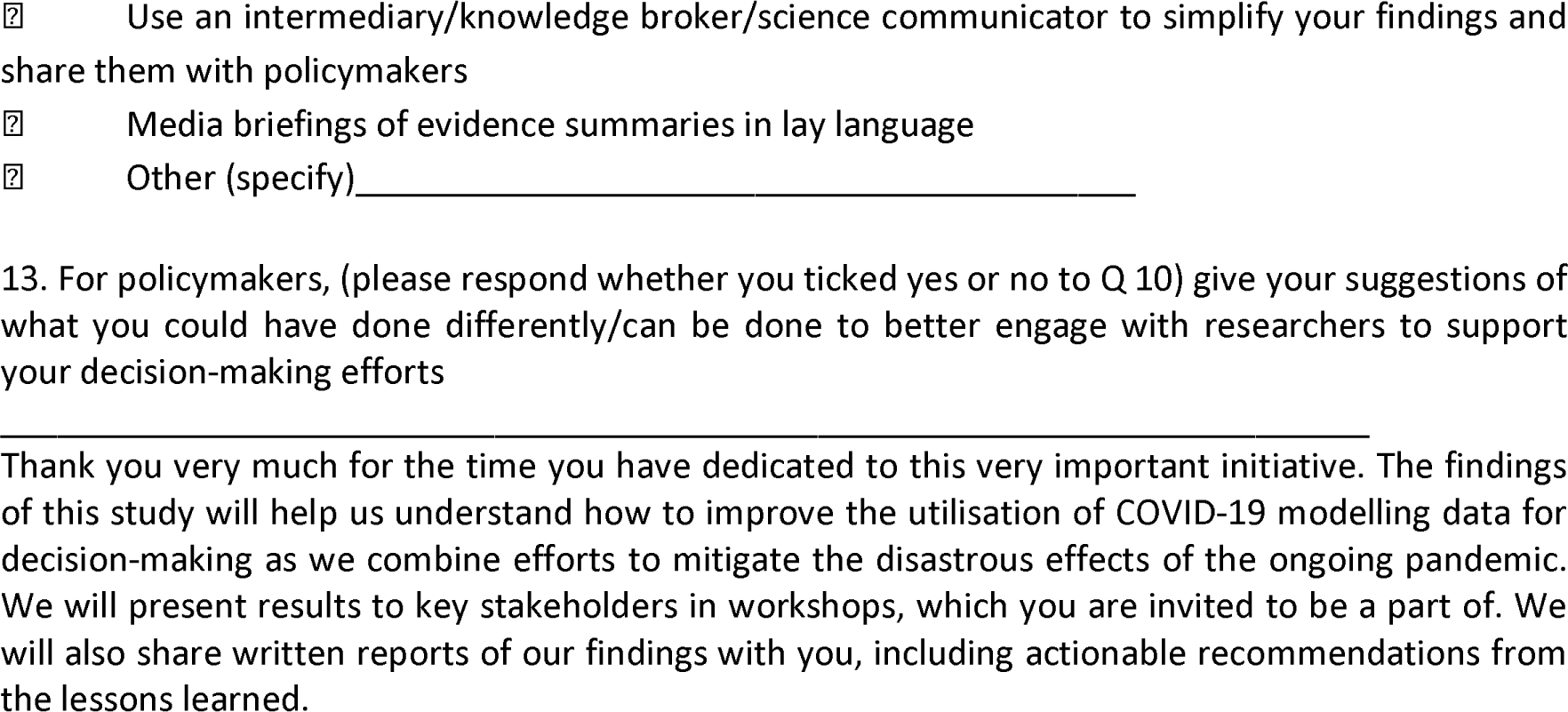

## S3 Scoping Review

### Specific objectives of the review

- Describe knowledge translation strategies and tools used in translating modelling evidence during the pandemic.
- Identifying barriers and facilitators to using knowledge translation strategies intended to promote uptake of COVID-19 modelling evidence for decision-making.
- Identify outcomes reported for those knowledge translation approaches.
- Identify key players in the COVID-19 modelling knowledge translation space.

### Search Strategy

The following search strategy was used:

Table: Search strings used in the scoping review. Concepts were combined using the Boolean operator “AND”

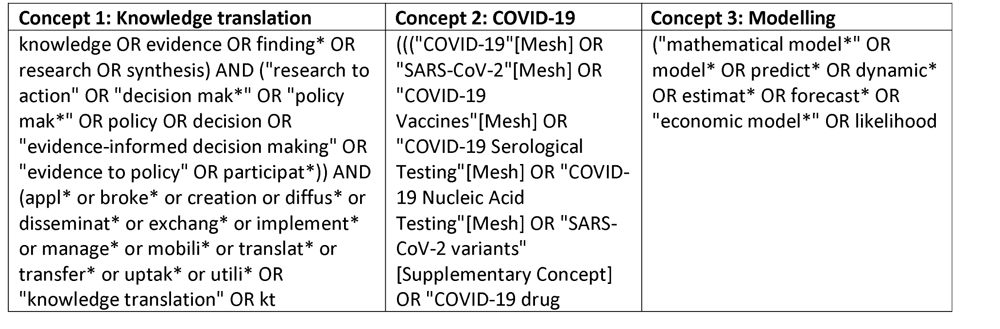
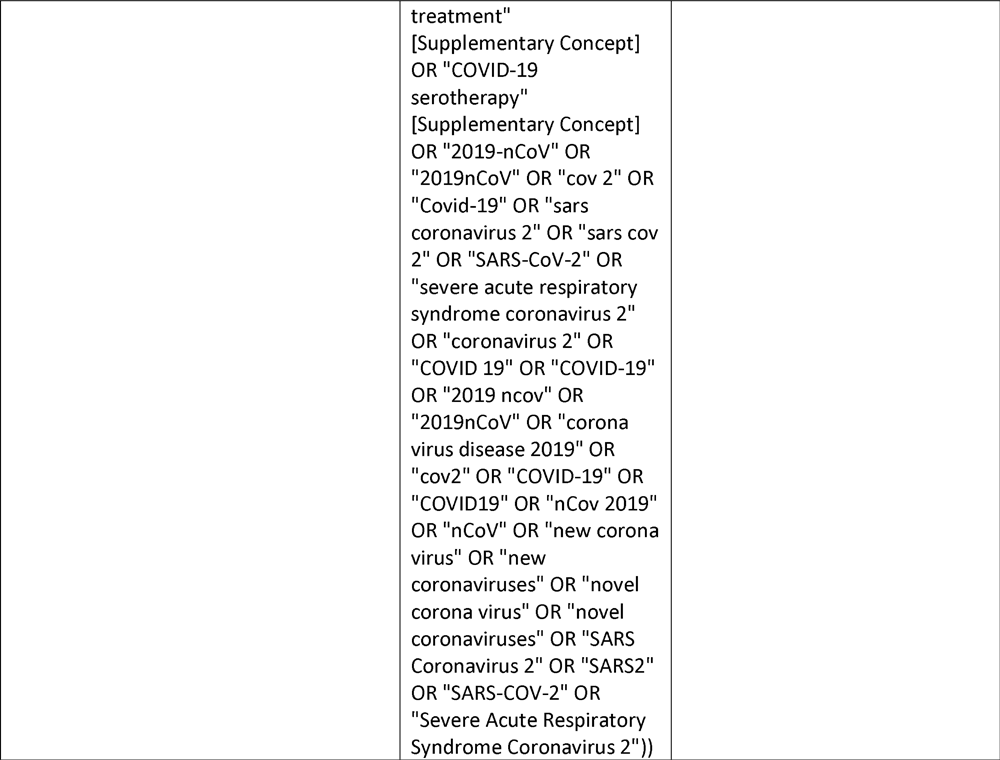

### Selection process

**Figure 1.**
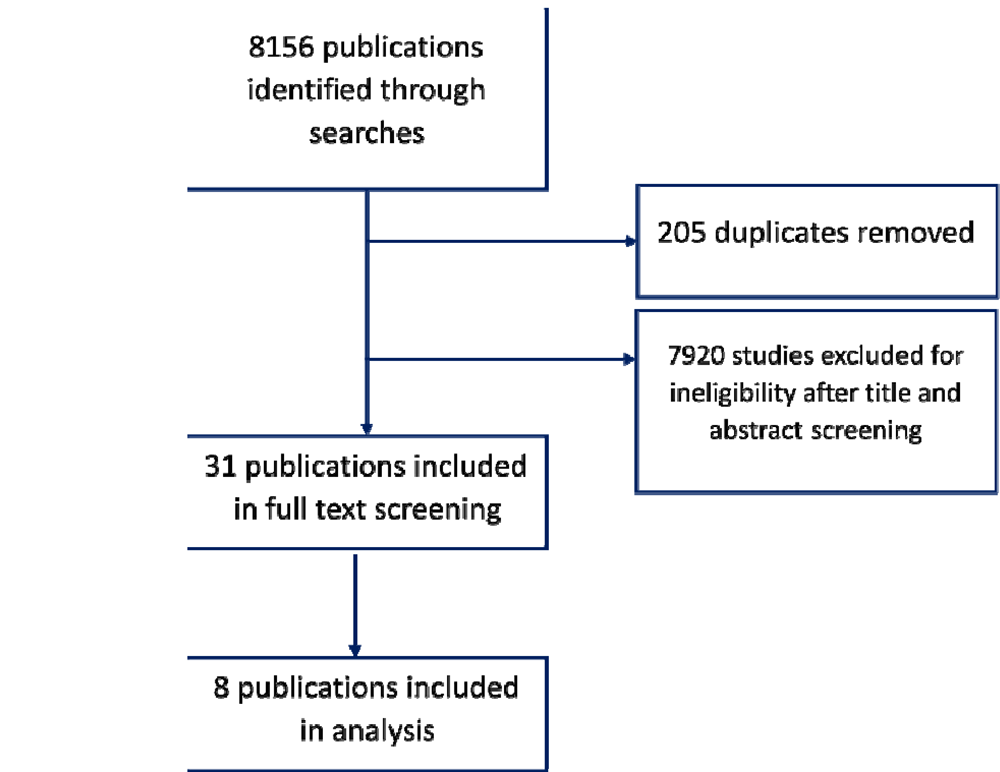
Selection process for the scoping review

### Table Scoping review: Characteristics of included studies

| Author | Study | Region | Experiences/ Findings |
| --- | --- | --- | --- |
| Adib et al.2021<sup>15</sup> | A participatory modelling approach for investigating the spread of COVID-19 in countries of the Eastern Mediterranean Region to support public health decision-making | Mediterranean | Limited surveillance data<br>Designed a participatory process to develop rapid modelling analyses tailored to the COVID-19 epidemic situation. |
| Abubakar et al.2021<sup>14</sup> | Lessons from co-production of evidence and policy in Nigeria's COVID-19 response | Africa - Nigeria | Limited transparency<br>Bureaucratic obstacles<br>Focus on epidemiological approaches |
| Sombie et al, 2020<sup>16</sup> | How does the West African Health Organisation (WAHO) contribute to the evidence-based decision making and practice during COVID-19 pandemic in ECOWAS region? | Africa – West African region | Capacity building to policy actors on the synthesis of recent data, development of guides and policy briefs |
| Hendy et al, 2021<sup>31</sup> | Mathematical modelling to inform New Zealand's COVID-19 response | New Zealand | Describes how the Stochastic model was used to compare the effects of various interventions in reducing the spread of the virus and to estimate the probability of elimination |
| Hillmer et al, 2021<sup>33</sup> | Ontario's COVID-19 Modelling Consensus Table: mobilising scientific expertise to support pandemic response | Ontario – East Central Canada | COVID-19 Modelling Consensus Table (MCT) is a partnership between the province and academic modellers and consists of multiple experts, health system leaders, and senior decision-makers. |
| Aguas et al., 2020<sup>27</sup> | Modelling the COVID-19 pandemic in context: an international participatory approach | Several | Describes the creation of a participatory modelling approach platform, the COVID-19 Modelling (CoMo) Consortium model, and illustrates some of its use cases |
| Teerawattananon Et al.,2022<sup>34</sup> | Recalibrating the notion of modelling for policymaking during pandemics | Several | Use fitness-for-purpose flowchart and reporting standards trajectory to address the challenges in using models for policymaking.<br>Adopting such tools can provide a strong justification for increased funding essential for preventing and responding to public health emergencies. |
| Gombos et al., 2020<sup>32</sup> | Translating Scientific Knowledge to Government Decision Makers Has Crucial Importance in the Management of the COVID-19 Pandemic | Hungary | Formation of a multidisciplinary research team performed a large amount of scientific data analysis and mathematical and socioeconomic modelling of the COVID-19 epidemic |

#### S4 Interview guides

#### Interview guide for researchers (English)

|  |
| --- |
| Interviewee code |
| Country code |
| Organisation code |
| Date |
| Position/Role |
| Age group |
| Gender |
| Period I have worked in this role. |
| Involvement in Technical Working Group (TWG) |

(After introducing yourself, explaining the purpose of the interview, seeking consent, and finding a setting and time that will make the interviewee comfortable, below is a list of topics to be discussed. The topic guide will remain flexible concerning what is relevant to participants)

- Can you tell me what your role(s) have been as an epidemiologist/statistician/modeler in this organisation, how long have you served in this capacity? Have your responsibilities changed in response to the ongoing COVID-19 pandemic? If yes, in what ways? How has this affected your morale/motivation? (Probe: type of modelling done: mathematical, economic, etc.; specify which disease areas they have worked in; identify if roles have changed due to demand from the pandemic?)
- What does it mean for you to be a scientist working in the public eye on COVID-19? (Probe: has the experience been positive or negative? In what ways?)
- Have you been involved in presenting evidence to inform policy? How has your experience in being part of this? (Probe: in what capacity? policy briefs; TWG meetings; news briefings; other?)
- What institutional/group changes, if any, have you had to make to accommodate evidence demands/support knowledge sharing?
- How have you found collaborating with government and policy organisations? How have these collaborations evolved, especially with the ongoing pandemic? (Probe: What impact has this experience had on your relationship with policymakers?)
- Have you been part of collaborative efforts with other modelling groups? Which ones? How has the experience been?
- Are you aware of any of the formal knowledge translation methods? Have you ever used any of them? Specify which? (Probe: -targeted dissemination; involving users in the research process; developing networks between researchers and users; use of knowledge brokers) (Probe: what capacity needs have you experienced? Has your capacity to communicate evidence improved?)
- Describe which approaches you/your organisation use to share your research findings with policymakers: i) outside the pandemic; ii) during the pandemic? (Probe: long-form reports; short country reports; policy/issue briefs; interactive websites; webinars; newspapers; blogs; websites etc.) ask them to specify where a combination has been used.
- Have policymakers ever approached you/your organisation to request data, or do you approach them to share your findings? Have you been part of a technical working group/advisory board? (Probe: how has this changed with the pandemic? What has been your experience in being part of a TWG/advisory board?)
- What has been your experience trying to make your research findings of complex mathematical models more palatable to lay people? (Probe: have you had any specific training in science communication? Does your organisation offer this support? Has this changed with the pandemic)?
- How have you been involved in presenting scientific evidence to the public, and how have you found this? (Probes: How has your role in informing the public about COVID-19 evolved?)
- How do you feel about how the press covers your scientific contributions?
- How do you feel about sharing your scientific evidence on social media?
- How do you feel about how the government presents (your) scientific evidence to the public?
- What kind of reactions have you received from the public? Have you received disturbing reactions or threats? How did you deal with this?
- How has your involvement in COVID-19 research and advising policy/government affected your professional life?
- How do your COVID-19 roles and responsibilities impact your existing roles?
- How has your involvement in COVID-19 research and advising policy/government affected your personal life? How have you dealt with that?
- What have you learned from your experience of informing COVID policy? (Probe: What do you think are the most important lessons from this experience? What can be done to improve things?)
- If a new pandemic breaks out in the future, what should be done differently regarding sharing information and bringing scientists and policymakers together?
- Do you have any additional remarks?
- Is there something you think we didn’t cover relevant to this issue/topic?
- Is there someone else you think we should talk to? (Thank the participant for their time and find out if they are okay with being followed up later to clarify things and to share the overall findings and invite them to be part of the learning sessions.)

#### Interview guide for policymakers (English)

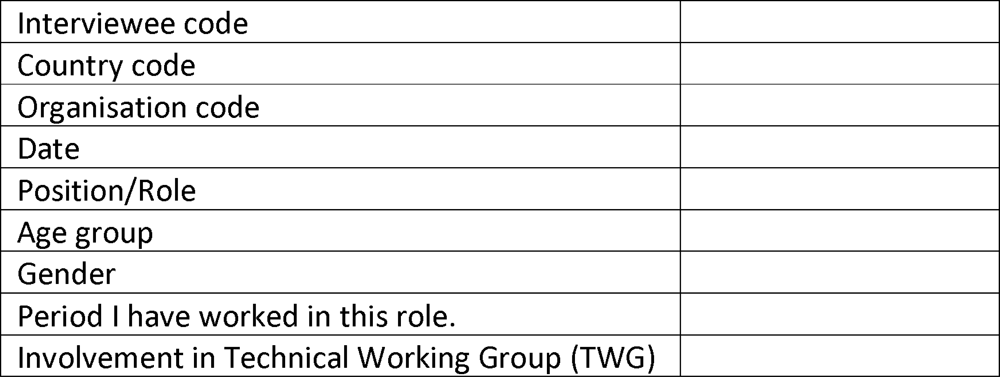

(After introducing yourself, explaining the purpose of the interview, seeking consent, and finding a setting and time that will make the interviewee comfortable, below is a list of topics to be discussed. The topic guide will remain flexible concerning what is relevant to participants)

- Can you tell me what your role(s) have been as a policy maker, how long have you served in this capacity? (Probes: Have your responsibilities changed in response to the ongoing COVID-19 pandemic? If yes, in what ways? How has this affected your morale/motivation?)
- What does it mean for you to be a policymaker working in the public eye on COVID-19? (Probe: has the experience been positive or negative? In what ways?)
- Have you been involved in using evidence to inform policy? How has your experience in being part of this? (Probe: in what capacity? Reading policy briefs/manuscripts; TWG meetings; news briefings; other?)
- How have you found collaborating with research organisations? How have these collaborations evolved, especially with the ongoing pandemic? (Probe: What impact has this experience had on your relationship with researchers?)
- Describe which approaches researchers use to share your research findings with you: i) outside the pandemic; ii) during the pandemic? (Probe: Probe: Have these approaches increased your accessibility to evidence?)
- Have you, as a policy maker, ever approached a researcher/research organisation to request data, or do they approach you to share their findings? Have you been part of a technical working group/advisory board? (Probe: how has this changed with the pandemic? What has been your experience in being part of a TWG/advisory board?)
- What has been your experience in trying to understand/make sense of research findings of complex mathematical models? (Probe: have you had any specific training in science communication? Does your institution offer this support? Has this changed with the pandemic? Has your capacity to use research evidence improved?)
- How have you assessed the quality of modelling evidence that you use to make decisions?
- Do you intend to continue seeking research evidence to make policy decisions in the future?
- In your opinion, what impact has evidence exchange had on COVID-19 policymaking? Give local examples if available.
- How have you been involved in presenting scientific evidence to the public, and how have you found this? (Probes: How has your role in informing the public about COVID-19 evolved?)
- How do you feel about how the press covers scientific contributions?
- How do you feel about sharing scientific evidence on social media?
- How do you feel about how scientific evidence is presented to the public?
- What kind of reactions have you received from the public? Have you received disturbing reactions or threats? How did you deal with this?
- How has your involvement in COVID-19 policy decision-making affected your professional life?
- How do your COVID-19 roles and responsibilities impact your existing roles?
- How has your involvement in COVID-19 policy decision-making affected your personal life? How have you dealt with that?
- What have you learned from your experience of generating COVID policies? (Probe: What do you think are the most important lessons from this experience? What can be done to improve things?)
- If a new pandemic breaks out in the future, what should be done differently regarding sharing information and bringing scientists and policymakers together?
- Do you have any additional remarks?
- Is there something you think we didn’t cover relevant to this issue/topic?
- Is there someone else you think we should talk to?

Thank the participant for their time, find out if they are okay with being followed up later to clarify things, share the overall findings, and invite them to be part of the learning sessions.

### S5 Coding framework

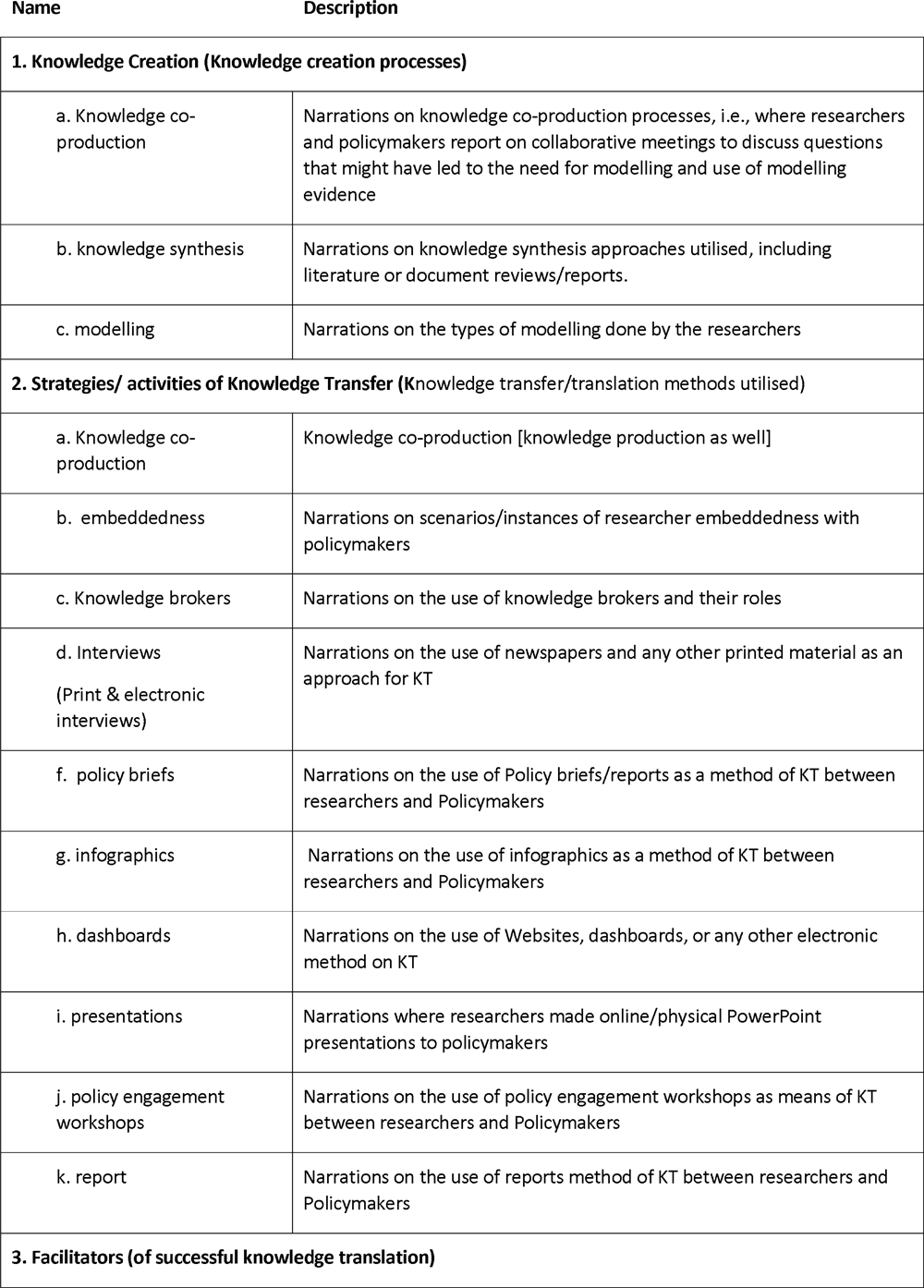
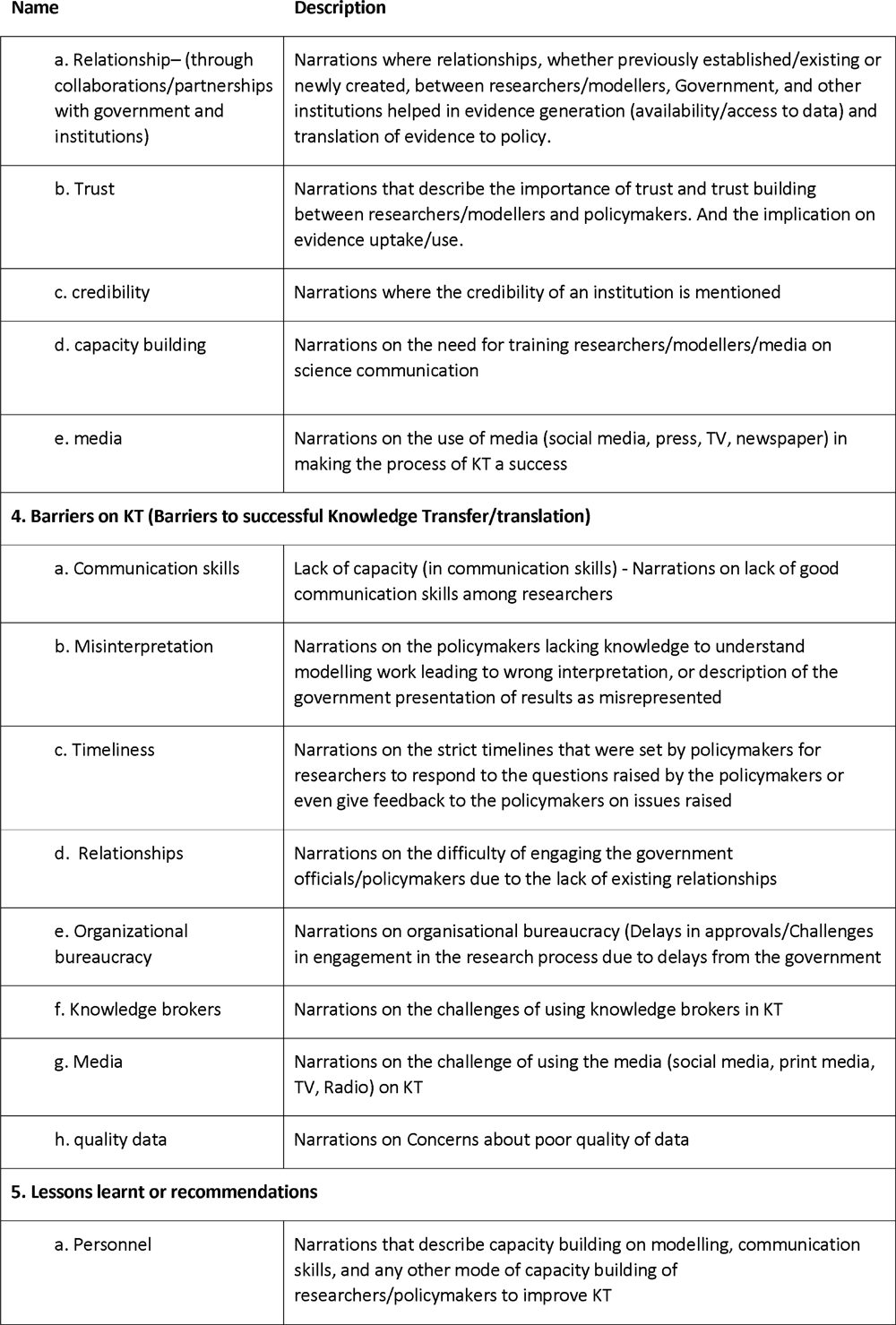
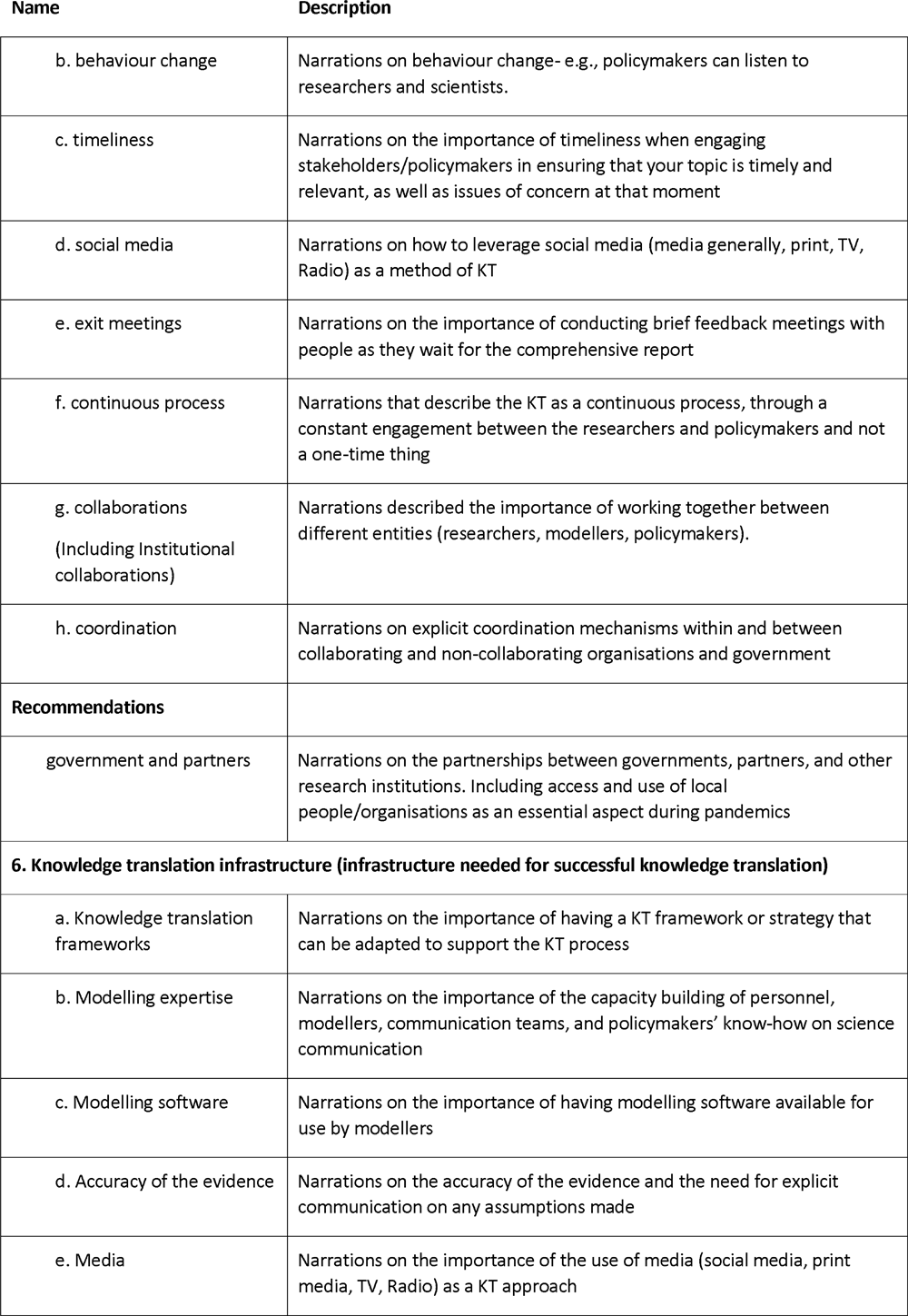
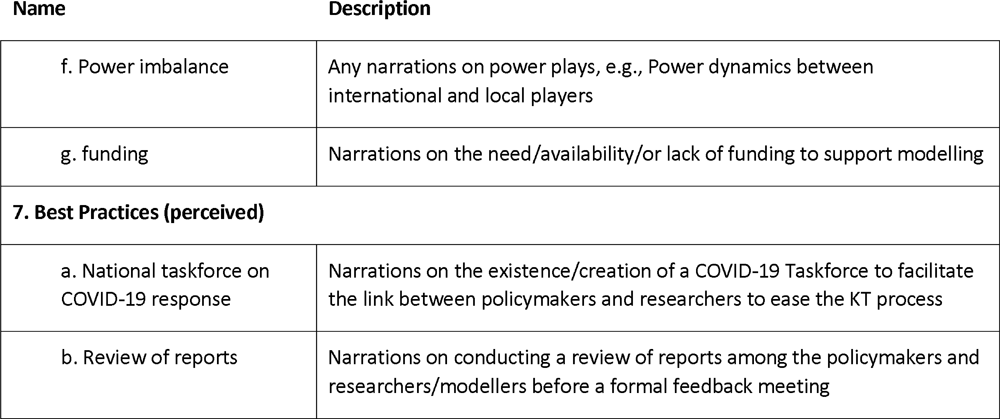

| Name | Description |
| --- | --- |
| **1. Knowledge Creation (Knowledge creation processes)** |  |
| a. Knowledge co-production | Narrations on knowledge co-production processes, i.e., where researchers and policymakers report on collaborative meetings to discuss questions that might have led to the need for modelling and use of modelling evidence |
| b. knowledge synthesis | Narrations on knowledge synthesis approaches utilised, including literature or document reviews/reports. |
| c. modelling | Narrations on the types of modelling done by the researchers |
| **2. Strategies/ activities of Knowledge Transfer (Knowledge transfer/translation methods utilised)** |  |
| a. Knowledge co-production | Knowledge co-production [knowledge production as well] |
| b. embeddedness | Narrations on scenarios/instances of researcher embeddedness with policymakers |
| c. Knowledge brokers | Narrations on the use of knowledge brokers and their roles |
| d. Interviews<br>(Print & electronic interviews) | Narrations on the use of newspapers and any other printed material as an approach for KT |
| f. policy briefs | Narrations on the use of Policy briefs/reports as a method of KT between researchers and Policymakers |
| g. infographics | Narrations on the use of infographics as a method of KT between researchers and Policymakers |
| h. dashboards | Narrations on the use of Websites, dashboards, or any other electronic method on KT |
| i. presentations | Narrations where researchers made online/physical PowerPoint presentations to policymakers |
| j. policy engagement workshops | Narrations on the use of policy engagement workshops as means of KT between researchers and Policymakers |
| k. report | Narrations on the use of reports method of KT between researchers and Policymakers |
| **3. Facilitators (of successful knowledge translation)** |  |
| a. Relationship– (through collaborations/partnerships with government and institutions) | Narrations where relationships, whether previously established/existing or newly created, between researchers/modellers, Government, and other institutions helped in evidence generation (availability/access to data) and translation of evidence to policy. |
| b. Trust | Narrations that describe the importance of trust and trust building between researchers/modellers and policymakers. And the implication on evidence uptake/use. |
| c. credibility | Narrations where the credibility of an institution is mentioned |
| d. capacity building | Narrations on the need for training researchers/modellers/media on science communication |
| e. media | Narrations on the use of media (social media, press, TV, newspaper) in making the process of KT a success |
| **4. Barriers on KT (Barriers to successful Knowledge Transfer/translation)** |  |
| a. Communication skills | Lack of capacity (in communication skills) - Narrations on lack of good communication skills among researchers |
| b. Misinterpretation | Narrations on the policymakers lacking knowledge to understand modelling work leading to wrong interpretation, or description of the government presentation of results as misrepresented |
| c. Timeliness | Narrations on the strict timelines that were set by policymakers for researchers to respond to the questions raised by the policymakers or even give feedback to the policymakers on issues raised |
| d. Relationships | Narrations on the difficulty of engaging the government officials/policymakers due to the lack of existing relationships |
| e. Organizational bureaucracy | Narrations on organisational bureaucracy (Delays in approvals/Challenges in engagement in the research process due to delays from the government |
| f. Knowledge brokers | Narrations on the challenges of using knowledge brokers in KT |
| g. Media | Narrations on the challenge of using the media (social media, print media, TV, Radio) on KT |
| h. quality data | Narrations on Concerns about poor quality of data |
| **5. Lessons learnt or recommendations** |  |
| a. Personnel | Narrations that describe capacity building on modelling, communication skills, and any other mode of capacity building of researchers/policymakers to improve KT |
| b. behaviour change | Narrations on behaviour change- e.g., policymakers can listen to researchers and scientists. |
| c. timeliness | Narrations on the importance of timeliness when engaging stakeholders/policymakers in ensuring that your topic is timely and relevant, as well as issues of concern at that moment |
| d. social media | Narrations on how to leverage social media (media generally, print, TV, Radio) as a method of KT |
| e. exit meetings | Narrations on the importance of conducting brief feedback meetings with people as they wait for the comprehensive report |
| f. continuous process | Narrations that describe the KT as a continuous process, through a constant engagement between the researchers and policymakers and not a one-time thing |
| g. collaborations<br>(Including Institutional collaborations) | Narrations described the importance of working together between different entities (researchers, modellers, policymakers). |
| h. coordination | Narrations on explicit coordination mechanisms within and between collaborating and non-collaborating organisations and government |
| **Recommendations** |  |
| government and partners | Narrations on the partnerships between governments, partners, and other research institutions. Including access and use of local people/organisations as an essential aspect during pandemics |
| **6. Knowledge translation infrastructure (infrastructure needed for successful knowledge translation)** |  |
| a. Knowledge translation frameworks | Narrations on the importance of having a KT framework or strategy that can be adapted to support the KT process |
| b. Modelling expertise | Narrations on the importance of the capacity building of personnel, modellers, communication teams, and policymakers' know-how on science communication |
| c. Modelling software | Narrations on the importance of having modelling software available for use by modellers |
| d. Accuracy of the evidence | Narrations on the accuracy of the evidence and the need for explicit communication on any assumptions made |
| e. Media | Narrations on the importance of the use of media (social media, print media, TV, Radio) as a KT approach |
| f. Power imbalance | Any narrations on power plays, e.g., Power dynamics between international and local players |
| g. funding | Narrations on the need/availability/or lack of funding to support modelling |
| **7. Best Practices (perceived)** |  |
| a. National taskforce on COVID-19 response | Narrations on the existence/creation of a COVID-19 Taskforce to facilitate the link between policymakers and researchers to ease the KT process |
| b. Review of reports | Narrations on conducting a review of reports among the policymakers and researchers/modellers before a formal feedback meeting |

### S6 Snapshot of the Thematic Analytic Framework matrix

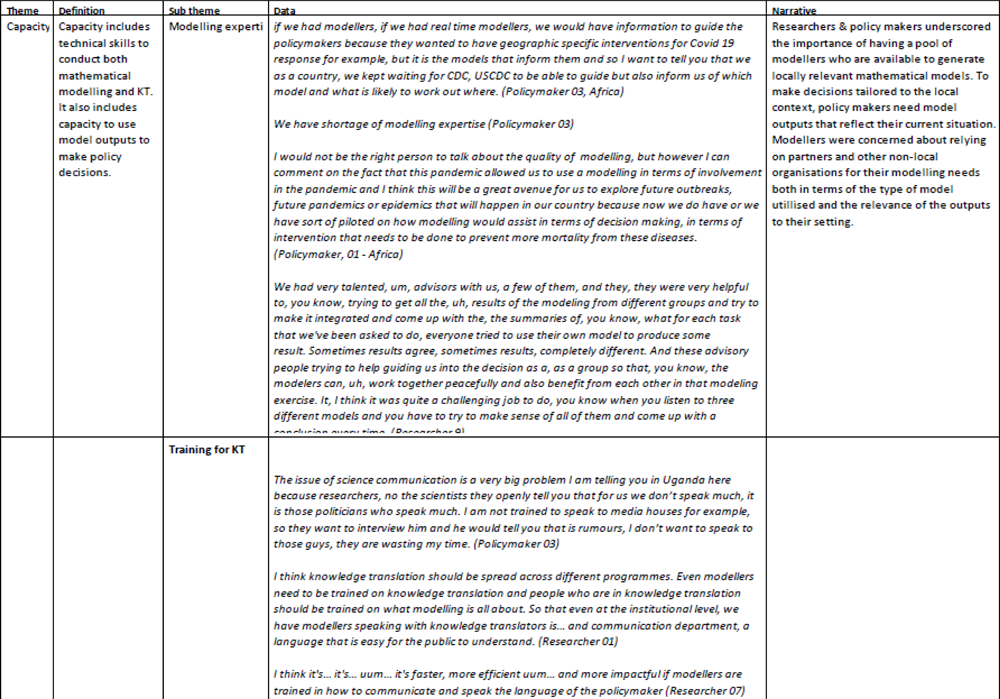

## Notes

### Competing Interest Statement

The authors have declared no competing interest.

